# PD-L1 expression is associated with higher residual cancer burden in triple-negative breast cancers with residual disease after neoadjuvant chemotherapy

**DOI:** 10.1101/2020.12.04.20244277

**Authors:** Beatriz Grandal, Manon Mangiardi-Veltin, Enora Laas, Marick Laé, Didier Meseure, Guillaume Bataillon, Elsy El-Alam, Lauren Darrigues, Elise Dumas, Eric Daoud, Anne Vincent-Salomon, Laure-Sophie Talagrand, Jean-Yves Pierga, Fabien Reyal, Anne-Sophie Hamy

**Affiliations:** Residual Tumor & Response to Treatment Laboratory, RT2Lab, Translational Research Department, INSERM, U932 Immunity and Cancer, University Paris, Paris, France; Department of Medical Oncology, Institut Curie, Paris, France; University Paris, Paris France; Department of Surgical Oncology, Institut Curie, Paris, France; University Paris, Paris France; Department of Pathology, Henri Becquerel Cancer Center, INSERM U1245; UniRouen Normandy University, Rouen, France; Department of Pathology, Institut Curie, Paris, France; University Paris, Paris France

**Keywords:** Breast cancer, neoadjuvant chemotherapy, PD-L1, immunohistochemistry, residual cancer burden, residual disease, immunotherapy, immune checkpoint inhibitor

## Abstract

The consequences of neoadjuvant chemotherapy (NAC) for PD-L1 activity in triple-negative breast cancers (TNBC) are not well understood. This is an important issue as immune checkpoint inhibitors (ICI) are undergoing rapid development and could be beneficial in patients who do not achieve a pathological complete response. We used immunohistochemistry to assess PD-L1 expression (E1L3N clone, cutoff for positivity: ≥ 1%) on both tumor (PD-L1-TC) and immune cells (PD-L1-IC) from a cohort of surgical specimens of T1-T3NxM0 TNBCs treated with NAC. PD-L1-TC was detected in 17 cases (19.1%) and PD-L1-IC in 14 cases (15.7%). None of the baseline characteristics of the tumor or the patient were associated with PD-L1 positivity, except for pre-NAC stromal TIL levels, which were higher in post-NAC PD-L1-TC-positive than in negative tumors. PD-L1-TC were significantly associated with a higher residual cancer burden (*p*=0.035) and aggressive post-NAC tumor characteristics, whereas PD-L1-IC were not. PD-L1 expression was not associated with DFS (*p*=0.38) or OS (*p*=0.48), but high Ki67 levels after NAC were strongly associated with a poor prognosis (DFS *p*=0.0014 and OS *p*=0.001). A small subset of TNBC patients displaying PD-L1 expression in the context of an extensive post-NAC tumor burden could benefit from ICI treatment after standard NAC.

## Introduction

Breast cancer (BC) remains the most frequent and deadly cancer in women [1]. Triple-negative (TNBC) subtypes account for 10 to 20% of all BCs, are more aggressive than other subtypes and are associated with a poorer prognosis [2]. Historically, chemotherapy was the only viable systemic treatment option for both local and advanced TNBC. However, the arrival of poly-ADP ribose polymerase (PARP) inhibitors, antiandrogen therapy and immunotherapy is gradually opening up new prospects for treatment [3].

In the last decade, neoadjuvant chemotherapy (NAC) has become a standard of care for TNBC [4]. Pathological complete response (pCR) after NAC occurs in approximately 30 to 50% of cases and is associated with longer disease-free (DFS) and overall survival (OS) [5,6]. For patients not achieving a pCR, the amount of residual disease (RD) can be assessed by determining the residual cancer burden (RCB) index, which can be used to classify patients into several prognostic groups [7,8]. The identification of patients with a poorer prognosis has important implications, as these patients may benefit from second-line treatments, such as adjuvant capecitabine [9,10].

Immune checkpoint inhibitor (ICI) therapies targeting PD-1 [11] and PD-L1 have been shown to be effective against cancers at various sites, and a number of blocking antibodies are currently being tested for use in BC [12,13]. Atezolizumab is used in combination with nab-paclitaxel for the routine treatment of advanced TNBC [14]. In the neoadjuvant setting, pCR rates were found to be higher in TNBC patients treated with durvalumab in addition to NAC than in the placebo arm (53.4% *versus* 44.2%, respectively) [15]. The emerging biomarkers for ICI efficacy include principally (i) programmed cell death-1 (PD1); (ii) its ligand-1 (PD-L1); and (iii) high levels of tumor-infiltrating lymphocytes (TILs) [16–18]. In the GeparNuevo trial, a trend towards higher pCR rates in tumors positive for PD-L1 at baseline was observed, which was significant for PD-L1-tumor cells in the durvalumab arm (*p*=0.045) and for PD-L1-immune cells in the placebo arm (*p*=0.040), suggesting that high levels of PD-L1 were linked to a stronger response but were not predictive of durvalumab response [15]. Other than these data for pre-NAC PD-L1 expression and the response to ICI, data for PD-L1 expression in tumor cells and/or immune cells in RD are scarce and the prognostic implications are unknown.

It has been suggested that chemotherapy could act as a functional immunotherapy by enhancing the release of tumor-associated antigens capable of triggering an immune response directed against tumor cells [24]. Taking this into account, experts have recommended the systematic assessment of TIL levels in the post-neoadjuvant setting [19]. As changes in the immune microenvironment following NAC could, theoretically, have a significant impact on PD-L1 expression, their assessment is of interest in the neoadjuvant setting [20,21]. We investigated the influence of NAC on TNBC tumors and their microimmune environment, by analyzing PD-L1 expression after NAC in 89 patients with RD after the treatment of TNBC with NAC. Estrogen receptor (ER), progesterone receptor (PR), and human epidermal growth factor receptor 2 (Her2) status and Ki67 levels were also assessed, to decipher the relationships between hormonal and proliferation pathways and PD-L1.

## Materials and Methods

### Patients and tumors

The analysis was performed on 89 patients with triple-negative, invasive, unilateral, non-recurrent, breast carcinoma, stage T1-T3NxM0 treated with NAC at Institut Curie, Paris, between 2002 and 2012 (NEOREP Cohort, CNIL declaration number 1547270). NAC regimens changed over the recruitment period (anthracycline-based regimen or sequential anthracycline-taxane regimens). Surgery was performed four to six weeks after the end of chemotherapy. The study was approved by the Breast Cancer Study Group of Institut Curie and was conducted according to institutional and ethical rules regarding research on tissue specimens and patients. Written informed consent from the patients was not required under French regulations.

### Tumor samples

Tumor samples were reviewed by two specialist pathologists (DM, ML). The pathological diagnosis was confirmed by initial core needle biopsy (CNB) before treatment. TNBCs were defined as tumors negative for ER, PR, and *HER2* expression. Cases were considered to be ER- or PR-negative if < 10% of the tumor cells expressed ER/PR, in accordance with the guidelines applied in France [22]. *HER2* expression was determined by immunohistochemistry (IHC), with scoring according to American Society of Clinical Oncology (ASCO)/College of American Pathologists (CAP) guidelines [23]. Tumor cellularity was defined as the percentage of tumor cells (*in situ* and invasive) in the specimen (biopsy or surgical specimen). Mitotic index was reported per 10 high-power fields (HPF) (1 HPF = 0.301 mm^2^).

We evaluated PD-L1 expression as a percentage of tumor cells by membrane staining (PD-L1-TC), and as a percentage of tumor-infiltrating lymphocytes (TILs) by membrane or cytoplasmic staining (PD-L1-IC; relative to total TILs). PD-L1 positivity was defined as expression by ≥1% of the cells, for both tumor and immune cells. For descriptive purposes, we also provide binned data, as follows: PD-L1-TC: 0, <1%, [2-24%], [25-50%] and >50% and PD-L1-IC: 0, <1%, [2-4%], [5-10%] and >10%. PD-L1-IC immunohistochemical expression was evaluated with the following primary antibodies: (i) ER with antibody: Diagomics 10045-10 mAb rabbit IgG SP1; (ii) PR with antibody: Leica NCL-L-PGR312 mAb mouse IgG 16; (iii) Her2 with antibody: Dako A0485 pAb rabbit IgG; (iv) Ki67 with antibody: Dako M7240 mAb mouse IgG1 MIB-1; (v) PD-L1 with antibody: Cell Signaling #13684S (E1L3N^®^) XP^®^ rabbit mAb.

### Survival endpoints

Relapse-free survival (RFS) was defined as the time from surgery to death, locoregional recurrence or distant recurrence, whichever occurred first, and overall survival (OS) was defined as the time from surgery to death. Patients for whom none of these events were recorded were censored at the date of last known contact. The cutoff date for survival analysis was February 1, 2019.

### Statistical analysis

The study population was described in terms of frequencies for qualitative variables, or medians and associated ranges for quantitative variables. Chi-squared tests were performed to search for differences between subgroups for each variable (considered significant for *p*-values ≤ 0.05). Continuous variables were compared between groups in Wilcoxon-Mann-Whitney tests for groups of fewer than 30 patients and for variables following multimodal distributions. Student’s *t*-tests were used in all other cases. Survival probabilities were estimated by the Kaplan–Meier method, and survival curves were compared in log-rank tests. Hazard ratios and their 95% confidence intervals were calculated with the Cox proportional hazards model. Variables with a *p*-value for the likelihood ratio test of 0.05 or lower in univariate analysis were selected for inclusion in the multivariate analysis. A forward stepwise selection procedure was used to establish the final multivariate model, with a significance threshold of 5%. Data were processed and statistical analyses were carried out with R software version 3.1.2 (www.cran.r-project.org, [14].

## Results

### Characteristics of the patients and tumors

In total, 1199 patients treated with NAC were included in the institutional cohort of the Institut Curie: 376 had a TNBC, and 231 had RD after completing NAC. The triple-negative tumors for which pCR was achieved were of higher grades, with higher levels of Ki67 and TILs than tumors for which pCR was not achieved (Table S1). We retrieved blocks for 122 patients with RD, 89 of which were reviewed and included in this analysis. The patients had a median age of 50.2 years old, most were premenopausal (*n*= 51, 57%) and had a normal BMI (*n*= 47, 52%). For 69.7% of the tumors, the diagnosis was made at the T2 stage (*n*=62), mostly with baseline axillary node involvement (*n*= 51, 57.3%). Primary treatment was an anthracycline-taxane regimen in 95% of the patients (Table 1).

**Table 1.** Patient characteristics among TNBC according to PD-L1-TC and PD-L1-IC expression

| Characteristics | Class | Overall | PDL-L1-TC |  | *p* | PDL-L1-IC |  | *p* |
| --- | --- | --- | --- | --- | --- | --- | --- | --- |
|  |  |  | Negative | Positive |  | Negative | Positive |  |
| n= |  | 89 | 58 | 31 |  | 59 | 30 |  |
| **Baseline characteristics** |  |  |  |  |  |  |  |  |
| Age |  | 50.20 [39.40, 57.90] | 49.03 (10.87) | 49.16 (11.23) | 0.960 | 48.94 (10.54) | 49.34 (11.85) | 0.871 |
| Family history | No | 68 (76.4) | 44 (75.9) | 24 (77.4) | 1.000 | 46 (78.0) | 22 (73.3) | 0.824 |
|  | Yes | 21 (23.6) | 14 (24.1) | 7 (22.6) |  | 13 (22.0) | 8 (26.7) |  |
| Menopausal status | Premenopausal | 51 (57.3) | 33 (56.9) | 18 (58.1) | 1.000 | 34 (57.6) | 17 (56.7) | 1.000 |
|  | Postmenopausal | 38 (42.7) | 25 (43.1) | 13 (41.9) |  | 25 (42.4) | 13 (43.3) |  |
| BMI classes | 18.5-24.9 | 47 (52.8) | 34 (58.6) | 13 (41.9) | 0.266 | 34 (57.6) | 13 (43.3) | 0.221 |
|  | <18.5 | 2 (2.2) | 2 (3.4) | 0 (0.0) |  | 2 (3.4) | 0 (0.0) |  |
|  | 25-29.9 | 22 (24.7) | 12 (20.7) | 10 (32.3) |  | 11 (18.6) | 11 (36.7) |  |
|  | ≥30 | 18 (20.2) | 10 (17.2) | 8 (25.8) |  | 12 (20.3) | 6 (20.0) |  |
| Smoking status | Never | 61 (71.8) | 42 (75.0) | 19 (65.5) | 0.452 | 42 (73.7) | 19 (67.9) | 0.366 |
|  | Current | 12 (14.1) | 8 (14.3) | 4 (13.8) |  | 9 (15.8) | 3 (10.7) |  |
|  | Former | 12 (14.1) | 6 (10.7) | 6 (20.7) |  | 6 (10.5) | 6 (21.4) |  |
| Comorbidity | No | 35 (45.5) | 25 (49.0) | 10 (38.5) | 0.524 | 27 (51.9) | 8 (32.0) | 0.162 |
|  | Yes | 42 (54.5) | 26 (51.0) | 16 (61.5) |  | 25 (48.1) | 17 (68.0) |  |
| Clinic T stage | T0-T1 | 6 (6.7) | 2 (3.4) | 4 (12.9) | 0.077 | 4 (6.8) | 2 (6.7) | 0.257 |
|  | T2 | 62 (69.7) | 39 (67.2) | 23 (74.2) |  | 38 (64.4) | 24 (80.0) |  |
|  | T3-T4 | 21 (23.6) | 17 (29.3) | 4 (12.9) |  | 17 (28.8) | 4 (13.3) |  |
| Clinical N stage | N0 | 38 (42.7) | 25 (43.1) | 13 (41.9) | 1.000 | 26 (44.1) | 12 (40.0) | 0.889 |
|  | N1-N2-N3 | 51 (57.3) | 33 (56.9) | 18 (58.1) |  | 33 (55.9) | 18 (60.0) |  |
| SBR grade | Grade I-II | 12 (13.8) | 11 (19.3) | 1 (3.3) | 0.084 | 10 (17.2) | 2 (6.9) | 0.323 |
|  | Grade III | 75 (86.2) | 46 (80.7) | 29 (96.7) |  | 48 (82.8) | 27 (93.1) |  |
| Mitotic index |  | 32.62 (25.80) | 23.00 [11.00, 40.25] | 35.50 [14.75, 52.75] | 0.165 | 24.00 [11.00, 41.00] | 30.00 [14.00, 52.00] | 0.283 |
| Mitotic index status | [0,20) | 34 (41.5) | 24 (44.4) | 10 (35.7) | 0.600 | 24 (43.6) | 10 (37.0) | 0.740 |
|  | ≥20 | 48 (58.5) | 30 (55.6) | 18 (64.3) |  | 31 (56.4) | 17 (63.0) |  |
| Ductal carcinoma in situ | No | 83 (93.3) | 53 (91.4) | 30 (96.8) | 0.601 | 55 (93.2) | 28 (93.3) | 1.000 |
|  | Yes | 6 (6.7) | 5 (8.6) | 1 (3.2) |  | 4 (6.8) | 2 (6.7) |  |
| % stromal lymphocytes |  | 25.00 [10.00, 40.00] | 20.00 [10.00, 30.00] | 30.00 [16.25, 40.00] | 0.053 | 25.00 [10.00, 32.50] | 25.00 [5.00, 40.00] | 0.823 |
| % intra-tumoral lymphocytes |  | 5.00 [1.00, 15.00] | 5.00 [2.00, 5.00] | 5.00 [1.00, 15.00] | 0.345 | 5.00 [2.00, 12.50] | 5.00 [1.00, 15.00] | 0.803 |
| BC surgery | Lumpectomy | 59 (66.3) | 41 (70.7) | 18 (58.1) | 0.334 | 42 (71.2) | 17 (56.7) | 0.257 |
|  | Mastectomy | 30 (33.7) | 17 (29.3) | 13 (41.9) |  | 17 (28.8) | 13 (43.3) |  |
| NAC regimen | anthra-taxans | 85 (95.5) | 55 (94.8) | 30 (96.8) | 0.761 | 56 (94.9) | 29 (96.7) | 0.773 |
|  | anthra | 3 (3.4) | 2 (3.4) | 1 (3.2) |  | 2 (3.4) | 1 (3.3) |  |
|  | taxanes | 1 (1.1) | 1 (1.7) | 0 (0.0) |  | 1 (1.7) | 0 (0.0) |  |
| **Post-NAC characteristics** |  |  |  |  |  |  |  |  |
| ypN stage | 0 | 53 (59.6) | 35 (60.3) | 18 (58.1) | 0.109 | 33 (55.9) | 20 (66.7) | 0.065 |
|  | [1-3] | 22 (24.7) | 17 (29.3) | 5 (16.1) |  | 19 (32.2) | 3 (10.0) |  |
|  | [4-9] | 13 (14.6) | 5 (8.6) | 8 (25.8) |  | 6 (10.2) | 7 (23.3) |  |
|  | 10 and more | 1 (1.1) | 1 (1.7) | 0 (0.0) |  | 1 (1.7) | 0 (0.0) |  |
| RCB index |  | 2.12 [1.54, 2.98] | 2.03 [1.45, 2.56] | 2.27 [1.67, 3.72] | 0.035 | 2.19 [1.52, 2.76] | 2.10 [1.63, 3.45] | 0.456 |
| RCB classes | RCB-I | 15 (17.2) | 12 (21.1) | 3 (10.0) | 0.040 | 13 (22.0) | 2 (7.1) | 0.185 |
|  | RCB-II | 53 (60.9) | 37 (64.9) | 16 (53.3) |  | 35 (59.3) | 18 (64.3) |  |
|  | RCB-III | 19 (21.8) | 8 (14.0) | 11 (36.7) |  | 11 (18.6) | 8 (28.6) |  |
| Ki67 status | [0,20) | 51 (57.3) | 34 (58.6) | 17 (54.8) | 0.905 | 34 (57.6) | 17 (56.7) | 1.000 |
|  | ≥20 | 38 (42.7) | 24 (41.4) | 14 (45.2) |  | 25 (42.4) | 13 (43.3) |  |
| Mitotic index |  | 31.28 (40.93) | 15.00 [0.00, 47.00] | 25.00 [3.00, 70.00] | 0.036 | 17.00 [0.00, 50.00] | 14.00 [2.50, 51.50] | 0.541 |
| Mitotic index status | [0,20) | 43 (55.1) | 29 (55.8) | 14 (53.8) | 1.000 | 28 (53.8) | 15 (57.7) | 0.936 |
|  | ≥20 | 35 (44.9) | 23 (44.2) | 12 (46.2) |  | 24 (46.2) | 11 (42.3) |  |
| % stromal lymphocytes |  | 15.00 [10.00, 30.00] | 15.00 [10.00, 30.00] | 20.00 [6.25, 30.00] | 0.860 | 15.00 [10.00, 30.00] | 22.50 [5.00, 30.00] | 0.850 |
| Lymphovascular invasion | No | 54 (70.1) | 32 (65.3) | 22 (78.6) | 0.335 | 32 (65.3) | 22 (78.6) | 0.335 |
|  | Yes | 23 (29.9) | 17 (34.7) | 6 (21.4) |  | 17 (34.7) | 6 (21.4) |  |

Missing data: TC= tumor cells; IC= immune cells; Smoking status, n=4; Comorbidity, n=12; SBR grade, n=2; Ki67 classes, n=81; Mitotic index class, n=7; % stromal lymphocytes, n=2; % intra-tumoral lymphocytes, n=2; Lymphovascular invasion, n=86; Lymphovascular invasion postNAC, n=12; RCB index, n=2; RCB classes, n=2; % stromal lymphocytes postNAC, n=2; % intra-tumoral lymphocytes postNAC, n=7. NB = Pre-NAC Ki67 not displayed in the table due to the high amount of missing values (n=81). Among the patients with pre-NAC Ki67 available, all values were 20% or higher.
Abbreviations: BMI= body mass index; T= tumor; N= node; SBR= grade Scarff-Bloom and Richardson; NAC=neoadjuvant chemotherapy; anthra-taxans= anthracyclines and taxanes; anthra= anthracyclines; RCB= Residual Cancer Burden.
The “n” denotes the number of patients. In case of categorical variables, percentages are expressed between brackets. In case of continuous variables, mean value is reported. In case of nonnormal continuous variables, median value is reported, with interquartile range between brackets.

All residual tumors remained negative for ER and PR after NAC (*n*= 89). Similarly, *HER2* expression was negative in 88 of 89 cases (scored 1+ in 3 patients, 0 in 85 patients), and 2+ in one patient (FISH-negative). No post-NAC switch in BC subtype was found in any of the 89 patients.

### Association between post-NAC PD-L1 expression and baseline clinical and pathologic patterns

PD-L1 was not expressed (0%) or was very weakly expressed (=<1%) in 81% (*n*= 72) of tumors and 84% (*n*= 75) of immune cells (Figs. 1A-B). PD-L1-TC was associated with PD-L1-IC expression (*p*< 0.001) (Figs. 1C-D). None of the baseline characteristics of the patients were significantly associated with post-NAC PD-L1-TC. High pre-NAC stromal (str) TIL levels were the only tumor characteristic associated with PD-L1 positivity in tumor cells (*p*=0.05), but no association was found in immune cells (Fig. 2, Table 1). The differences in str TIL levels before and after NAC were not associated with PD-L1-TC nor PD-L1-IC positivity (Fig. S1).

**Figure 1.**
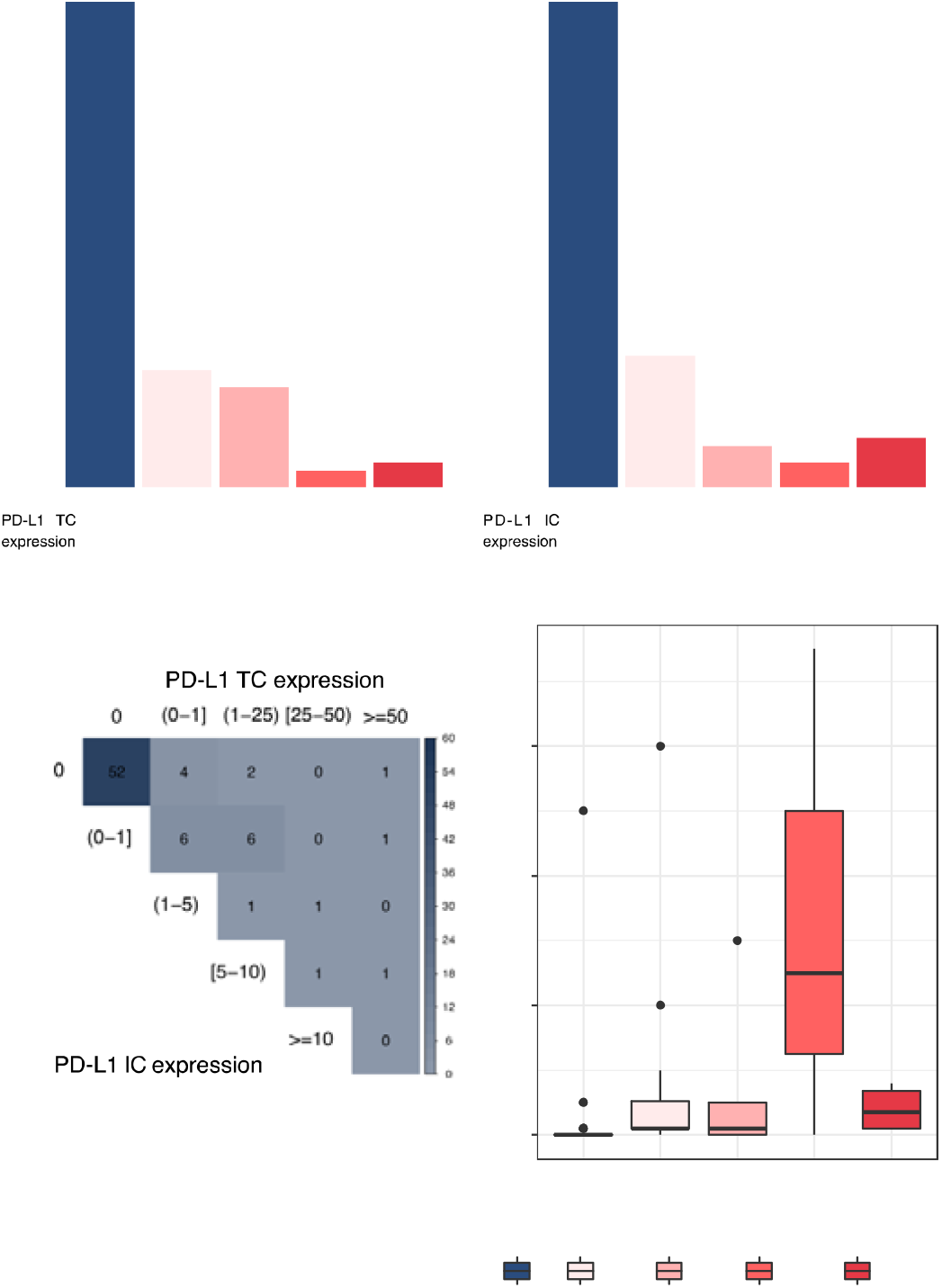
PD-L1-TC & PD-L1-IC expression after NAC. **A**, Barplot of distribution of PD-L1 tumor cells. **B**, Barplot of distribution of PD-L1 immune cells. **C**, Correlation between PD-L1 tumor cells and PD-L1 immune cells expression. **D**, PD-L1 tumor cells expression (continuous variable) by PD-L1 immune cells expression.

**Figure 2.**
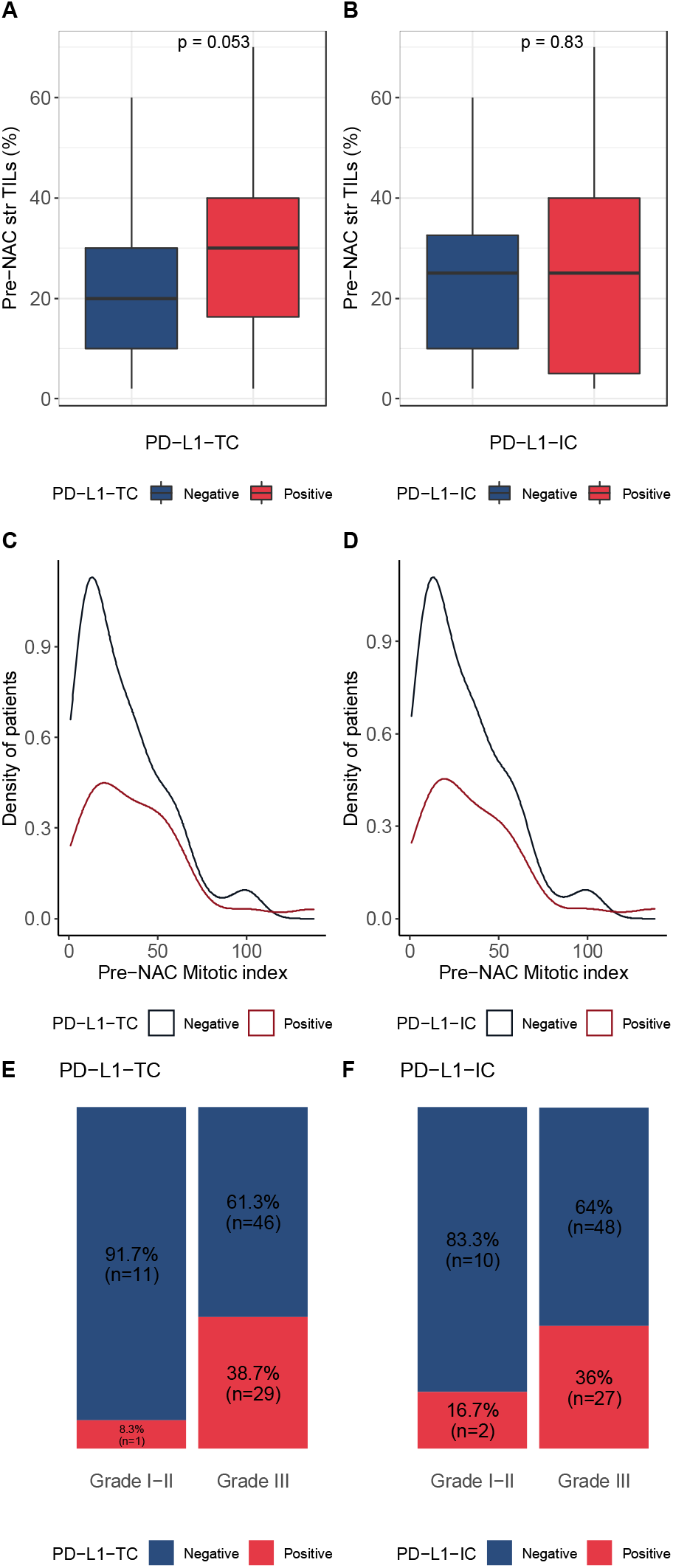
Baseline characteristics by PD-L1-TC & PD-L1-IC expression. Bottom and top bars of the boxplots represent the first and third quartiles, respectively, the medium bar is the median, and whiskers extend to 1.5 times the interquartile range. **A**, Associations between pre-NAC TILs and PD-L1-TC. **B**, Associations between pre-NAC TILs and PD-L1-IC. **C**, Kernel density plot of pre-NAC mitotic index in PD-LI-TC. **D**, Kernel density plot of pre-NAC mitotic index in PD-L1-IC. **E**, Percentage of tumor according to grade by PD-L1-TC. **F**, Percentage of tumor according to grade by PD-L1-IC.

### Association between post-NAC PD-L1 expression, KI67, and post-NAC pathological patterns

RCB was assessed in 87 of 89 tumors (98%). The proportion of patients within each RCB class was as follows: RCB-I, *n*=15 (17%); RCB-II, *n*=53 (61%), and RCB-III, *n*=19 (22%). PD-L1 expression was significantly associated with a higher RCB, higher post-NAC mitotic index, and a trend towards higher tumor cellularity (p=0.06), in tumor cells but not in immune cells (Fig. 3A-K). PD-L1-TC expression was not associated with Ki67 expression, but Ki67 was associated with RCB index (Fig. 3L-N).

**Figure 3.**
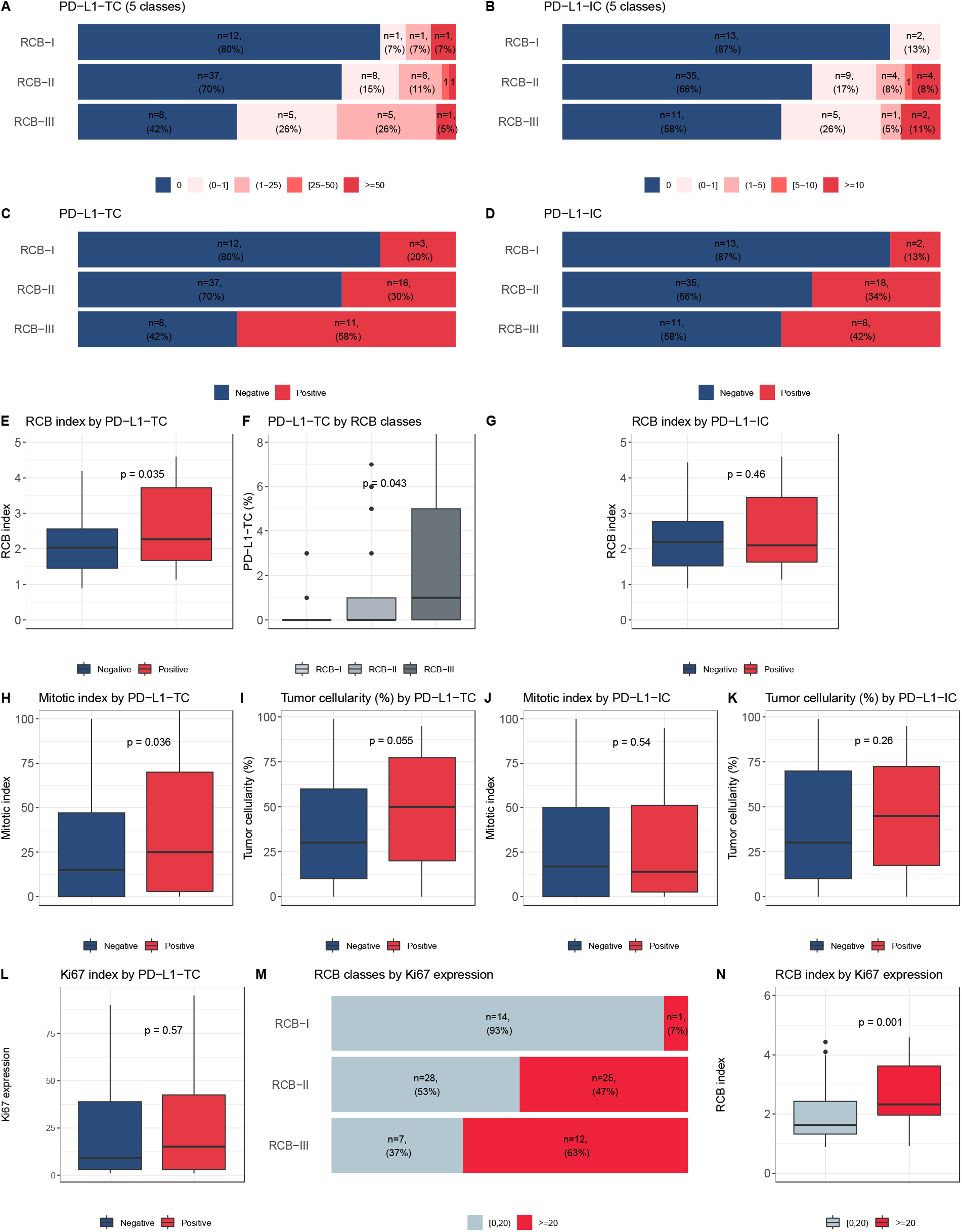
Post-NAC characteristic by PDL1 expression. Bottom and top bars of the boxplots represent the first and third quartiles, respectively, the medium bar is the median, and whiskers extend to 1.5 times the interquartile range. A, Percentage of tumor according to RCB classes by PD-L1-TC (5 classes). B, Percentage of tumor according to RCB classes by PD-L1-IC (5 classes). **C**, Percentage of tumor according to RCB classes by PD-L1-TC (2 classes). **D**, Percentage of tumor according to RCB classes by PD-L1-IC (2 classes). **E**, RCB-index by PD-L1-TC. **F**, PD-L1-TC by RCB classes. **G**, RCB-index by PD-L1-IC. **H**, Mitotic index by PD-L1-TC. **I**, Tumor cellularity (%) by PD-L1-TC. **J**, Mitotic index by PD-L1-TC. **K**, Tumor cellularity (%) by PD-L1-TC.

### Survival analyses

With a median follow-up of 80 months, 34 patients experienced relapses and 30 patients died. Neither PD-L1-TC nor PD-L1-IC was significantly associated with RFS or OS (Fig. 4A-D). By contrast, both RCB and Ki67 were significantly associated with RFS and OS (Fig. 4E-H), and were independent predictors of survival in multivariate analysis (RCB index, OR = 1.7; CI95% [1.25–2.42], *p<* 0.001; post-NAC Ki67 expression, OR = 2.7; CI95% [1.29–5.8], *p=* 0.008, respectively).

**Figure 4.**
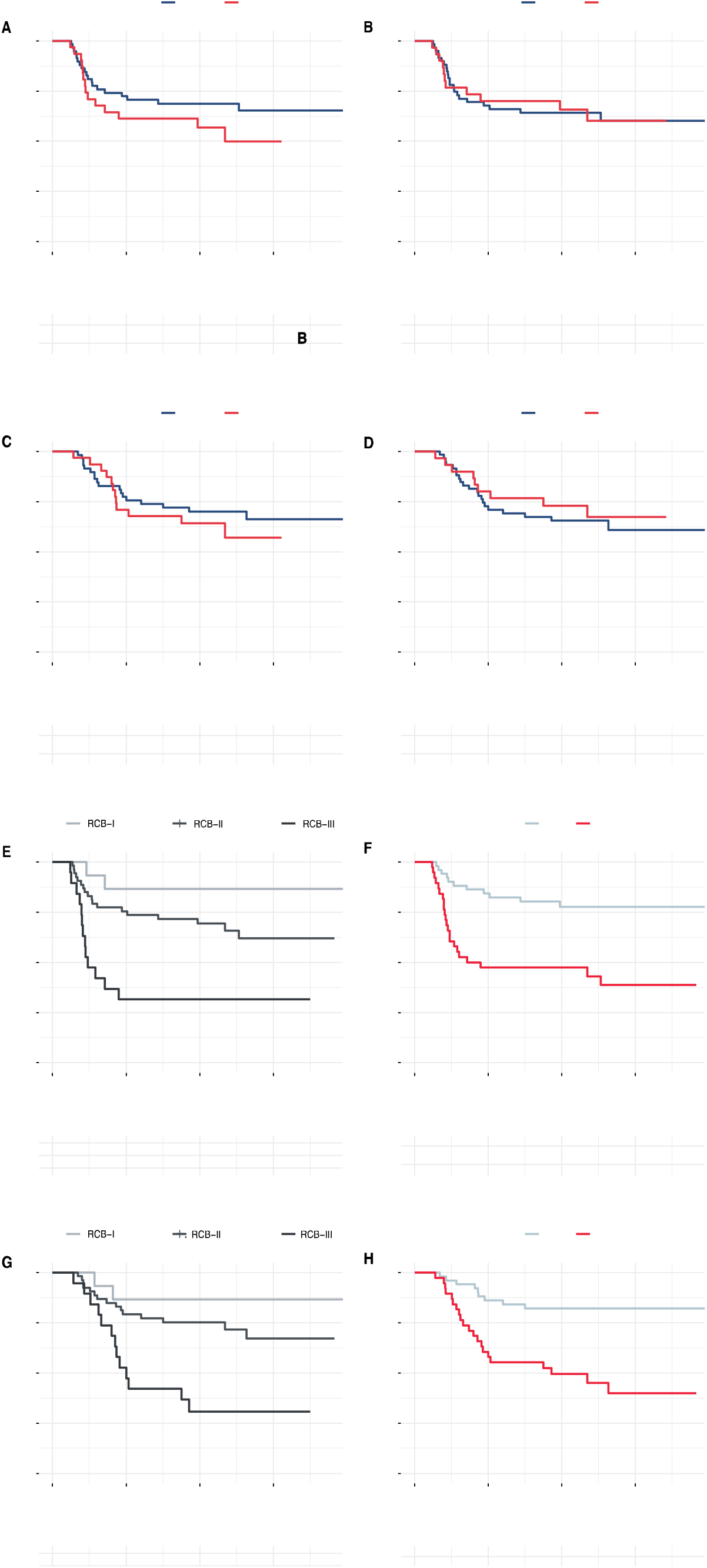
Survival curves. **A**, Relapse free survival curves according to PD-L1-TC expression. **B**, Relapse free survival curves according to PD-L1-IC expression. **C**, Overall survival curves according to PD-L1-TC expression. **D**, Overall survival curves according to PD-L1-IC expression. **E**, Relapse free survival curves according to RCB classes. **F**, Relapse free survival curves according to Ki67 expression. **G**, Overall survival curves according to RCB classes. **H**, Overall survival curves according to Ki67 expression.

## Discussion

In this study, we analyzed PD-L1 expression in tumor and immune cells from residual post-NAC TNBC. We found that PD-L1 expression was significantly associated with a greater tumor burden and aggressive post-NAC tumor characteristics. This study provides new insight into immunity in the post-NAC setting.

*First*, we observed that the proportions of tumor and immune cells positively stained for PD-L1 (19.1 and 15.7%, respectively) were low. None of the clinical or pathological features of the patients at baseline were associated with post-NAC PD-L1 expression. At least seven meta-analyses, including more than 125 studies, have evaluated PD-L1 expression in BC bulk tumor samples over the last three years (see Table S3). The prevalence of PD-L1 positivity ranged from 6.4 to 76.4% [24]. PD-L1 expression was significantly associated with the HR-negative [17,25,26], *HER2*-positive [25] and TNBC [17,25,26] subtypes, and the levels detected depended on the detection method [24]. To our knowledge, only three studies have analyzed PD-L1 expression on specimens of tumors previously treated with systemic therapy (see Table 2). In the SWOG N0800 neoadjuvant trials, Pelekanou *et al*. retrospectively analyzed 43 surgical specimens from *HER2*-negative tumors (TNBC, *n*=9) by assessing PD-L1 expression with the Dako clone and a 1% cutoff for positivity. They found that PD-L1 expression was relatively stable before and after treatment (43% (52/120) *versus* 33% (14/43), respectively) [27]. In a study on the same cohort of patients, Li *et al*. showed that PD-L1 levels in tumor and stromal cells were not significantly affected by treatment (*p*=0.502 and *p*=0.655, respectively) [28]. This cohort included both HR-positive and HR-negative BC specimens, and subgroup analyses were not possible due to the small sample size [28]. In a TNBC cohort including 114 patients, Wang *et al*. found that 37.7% of post-NAC specimens from patients with RDwere positive for PD-L1. This rate is higher than that for the cohort reported here, and such discrepancies between studies may be due to the use of a different antibody for PD-L1 expression (EPR19759 clone rather than the E1L3N clone used here).

**Table 2:** Studies on PD-L1 expression in BC RD

| First author | Journal | Year | Country | n patients (RD) | Detection technique (IHC clone) | Cut-off | PDL-1 prevalence in CNB | PDL1 and pCR | PDL-1 prevalence in RD | PDL1 and DFS | PDL1 and OS |
| --- | --- | --- | --- | --- | --- | --- | --- | --- | --- | --- | --- |
| Pelekanou (SWOG S0800) | Breast cancer research | 2017 | USA | 58 (Her2 negative) | RD: SP142 and 22C3 Dako combined CNB : E1L3N | QIF = 500 | 21/41 (51.0%) | p = 0.018 | 10 (17.2%) | - | - |
| Pelekanou (SWOG S0800) | Molecular cancer therapeutics | 2018 | USA | 43 (Her2 negative) out of which 9 TNBC | 22C3 Dako | QIF = 500 | 52/120 (43%) | higher pCR rates immune + tumor c. (p = 0.008) sole tumor c. NS (p = 0.10) | 14 (33%) | - (in CNB NS p=0.14) | - (in CNB NS p=0.64) |
| Wang | Journal of Breast Cancer | 2018 | China | 114 TNBC | EPR19759 | H-score of 100 | Tumor and/or Immune c. 48 (32.4) | HR = 1.34 [0.54; 3.30] p = 0.53 | 43 (37.7%) | Tumor/Immune c. NS ; HR = 1.421 [0.78; 2.59] p = 0.249 | - |
| Li (SWOG S0800) | Journal for Immunotherapy of Cancer | 2019 | USA | 60 (Her2 negative) | 22C3 Dako | 1% | - | NS in tumor c. (p= 0.1578) and immune c. (p=0.0722) | - | - | - |
| Loibl (GeparNuevo study) | Annals of Oncology | 2019 | Germany | 174 TNBC | SP263 Ventana | 1% | Tumor and/or Immune c. 138/158 (87.3) | Durvalumab group 47/88 (53.4%) Placebo group 38/86 (44.2%) | - | - | - |
| Srivastava | Journal of Family Medicine and Primary Care | 2020 | India | 30 | Abcam NA | H-score of 100 | 11 (36.7%) | - | 4 (13.3%) | - | - |
| Grandal | Cancers | 2020 | France | 89 TNBC | E1L3N | 1% | - | - | Tumor c. n = 17 (19.1%) Immune c. n = 14 (15.7%) | Tumor c. NS (p = 0.38) Immune c. NS (p = 0.23) | Tumor c. NS (p = 0.48) Immune c. NS (p = 0.46) |

*Abbreviations:* residual disease (RD), core needle biopsy (CNB), breast cancer (BC), cells (c.), triple-negative breast cancer (TNBC), hazard ratio (HR), confidence interval (CI), not significant (NS), relative risk (RR), pathological complete response (pCR)

*Second*, PD-L1 expression in post-NAC tumor cells was associated with a higher RCB index. High levels of PD-L1 in tumor cells have been linked to clinical and pathological features associated with a poor prognosis in numerous studies. In particular, an association with the presence of lymph node metastasis and high tumor grade has been reported for evaluations of non-pretreated tumors (see Table S3). In the pre-NAC setting, Huang *et al*. published a pooled analysis of eight studies including 1085 patients, and showed that PD-L1 positivity in core specimens before treatment was associated with higher pCR rates (RR = 1.64, _95%_CI [0.99; 2.73], *p* = 0.05), but with significant heterogeneity between studies (I^2^ = 79%, P_heterogeneity_ < 0.0001) [26]. No data have been published concerning the association between PD-L1 and response to treatment after NAC. It remains unknown whether the greater RCB observed here for PD-L1-positive tumors is a cause or consequence of the RD. Two mechanisms may be involved: the PD-L1 protein may operate as a suppressor of anti-tumor immune responses [29], preventing immune cells from clearing tumor cells efficiently, or chemotherapy-resistant tumor cells may be more likely to express immunosuppressive factors.

We found no impact of post-NAC PD-L1 expression on oncological outcomes, in terms of either DRFS or OS for either tumor cells or immune cells. Studies evaluating the prognostic implications of PD-L1 in BC have reported conflicting results. Some reported a positive association between PD-L1 and survival [30–34], whereas others found a negative association [17,25,26] or no association at all [35]. However, these findings must be interpreted with care, because PD-L1 expression may be differentially regulated on tumor or immune cells [29,36,37], and previous studies have highlighted the importance of studying the expression of this molecule in both types of cell [29]. In a recent large meta-analysis, Matikas and coworkers showed that PD-L1 expression in tumor cells was associated with a shorter DFS and OS, whereas PD-L1 expression in immune cells was associated with a longer DFS (HR = 0.61, _95%_CI [0.51–0.73], *p* < 0.001), and with a longer OS (HR = 0.53, _95%_CI [0.39–0.73], *p* < 0.001) in the TNBC subgroup (8 studies, *n*=969) [38]. These studies were performed on bulk tumors, and the prognostic value of PD-L1 may be different in the post-NAC setting. In the only study to date evaluating PD-L1 expression and prognosis in the context of RD, Wang and coworkers found no association between high levels of PD-L1 expression and DFS (*p*=0.249).

*Finally*, our study confirms the strong prognostic value of Ki67 expression after NAC, with high levels of expression associated with a poor prognosis. Post-NAC, high Ki67 levels have consistently found to be a predictor of poor DFS across studies [39–51]. In a matched cohort of 103 BC patients, Jones *et al*. found post-therapy Ki67 to be the only significant independent prognostic factor for DFS in multivariate analysis (*p*<0.001), and the strongest prognostic factor for OS (*p*<0.001) [49]. Ultimately, baseline Ki67 expression is predictive of pCR in patients with RD and persistent highly proliferative disease, but the outcome is poor [51]. The RCB index, first published in 2007, combines pathological findings from both the primary tumor bed and the regional lymph nodes. It has been validated as a predictor of the risk of recurrence in several independent cohorts [8,39] and is now widely used as a primary endpoint in clinical trials [52]. Several studies have suggested that the inclusion of additional factors, such as immunological features [53,54], lymphovascular invasion [55] or post-NAC Ki67 expression [56], could improve its performance. Our results confirm that amidst the candidate post-NAC parameters for improving RCB performance, post-NAC Ki67 expression, does indeed have a strong prognostic value, but our results are not consistent with the hypothesis that PD-L1 expression is a strong prognostic marker that could be used to refine estimates of the risk of relapse in the post-NAC setting, despite its association with greater RCB.

This study has several strengths and limitations. First, we provide unprecedented data on PD-L1 expression, using the E1L3N clone in the setting of post-NAC RD in TNBC patients. Our results call for additional evidence on the clinical relevance of such a biomarker for accurately identifying patients in whom it might be a useful theranostic marker. However, our cohort is potentially subject to technical pitfalls, as AFA fixative (a combination of alcohol, formalin, and acetic acid) was used in this historical cohort. It has recently been shown that the use of this fixative decreases antibody binding to PD-L1, and this has led to a switch to formol-only fixation [14]. Similar analyses on more recent paraffin-embedded specimens may therefore be of interest in this context. In addition, as in other studies, the main limitation of our report is the absence of a standardized detection technique and cutoff, as multiple assays and scoring systems exist. A wide range of positivity rates and discordant associations with prognosis have been reported across studies, depending on the antibodies used [26,38]. In our study, PD-L1 expression was assessed with the E1L3N antibody, which has been shown to be more sensitive than 28-8 Dako [57] and the SP142 Ventana Assay [58], but inferior to the SP263 assay [59,60]. Furthermore, different cutoffs are used to define PD-L1 positivity for different tumor types and in clinical trials [20]. The principal cutoffs used are 1 and 5% [38], but cutoffs values ranging from 1 to 50% have been used [26] and several teams use composite scores of staining intensity and the percentage of positive cells [26,38]. A standardized methodology is much needed, to ensure consistency and reproducibility in future PD-L1 studies [20]. Several teams have tried to overcome these discrepancies by using DNA microarrays to analyze PD-L1 mRNA levels [61–63].

Our study opens up several pragmatic perspectives. The TIL working group recently suggested that PD-L1 expression should be included in the routine clinical assessment of BC specimens [20]. Our results suggest that (i) further evidence is required to confirm the clinical utility of this marker and its validity in the post-NAC setting (ii) standardized guidelines for the assessment of this biomarker should be published before its integration into routine practice. Furthermore, as increasing numbers of patients with TNBC are being treated by NAC, the number of second-line trials in the post-NAC setting is growing [12,13]. Further studies are required to evaluate the proper place of PD-L1 as an immuno-oncological biomarker for selecting patients likely to benefit from ICI in the post-NAC setting.

In conclusion, our study identifies a small subset of patients with TNBCs and RD after NAC displaying PD-L1 expression in the context of a higher post-NAC tumor burden. As RCB is associated with a higher risk of relapse, and as these patients could theoretically respond to ICI [16], they could be invited to participate in second-line treatment trials of immunotherapy after NAC.

## Data Availability

The authors confirm that the data supporting the findings of this study are available within the article [and/or] its supplementary materials.

## Author Contributions

Conceptualization, A-S.H., L-S.T., M.L. and F.R.; methodology, A-S.H., L-S.T., M.L. and F.R.; software, A-S.H., B.G., E.L. and F.R.; validation, A-S.H., and F.R.; formal analysis, A-S.H. and B.G.; investigation, G.B, E.E-A., M.L, D.M., A.V-S.; resources, A-S.H. and F.R.; data curation, B.G. and A-S.H.; writing—original draft preparation, M.M-V., A-S.H.; writing—review and editing, M.M-V., B.G., A-S.H. and F.R..; visualization, A-S.H. and B.G.; supervision, A-S.H. and F.R.; project administration, F.R.; funding acquisition, A-S.H. All the authors have read and approved the version of this manuscript submitted.

