## Supplemental tables and figures for "PD-L1 expression is associated with higher residual cancer burden in triple-negative breast cancers with residual disease after neoadjuvant chemotherapy"

**SUPPLEMENTARY DATA**

**Supplementary tables**

**Table S1:** Patient characteristics among the whole population and by pCR status.

*Missing data:* Family history, n=1; Menopausal status, n=4; BMI classes, n=1; Smoking status, n=71; Comorbidity, n=56; Nuclear ER staining (%), n=20; Nuclear PR staining (%), n=18; SBR grade, n=9; KI67 classes, n=209; Mitotic index class, n=42; Ductal carcinoma in situ, n=43; % stromal lymphocytes, n=57; % intra-tumoral lymphocytes, n=57; Lymphovascular invasion, n=256; NAC regimen, n=1; Lymphovascular invasion postNAC, n=142; RCB index, n=57; RCB classes, n=57; % stromal lymphocytes postNAC, n=57; % intra-tumoral lymphocytes postNAC, n=197.

*Abbreviations*: BMI= body mass index; T= tumor; N= node; ER= estrogen receptor; PR= progesterone receptor; SBR= grade Scarff-Bloom and Richardson; BC= Breast cancer; NAC=neoadjuvant chemotherapy; anthra-taxans= anthracyclines and taxanes; anthra= anthracyclines; RCB= Residual Cancer Burden.

*The “n” denotes the number of patients. In case of categorical variables, percentages are expressed between brackets. In case of continuous variables, mean value is reported. In case of nonnormal continuous variables, median value is reported, with interquartile range between brackets.*

**Table S2:** Association of clinical and pathological pre and post-NAC parameters with PD-L1-TC expressio after univariate and multivariate analysis in the whole population.

*Abbreviations*: BMI=body mass index; ER=oestrogen receptor; PR=progesterone receptor; TNBC= triple negative breast cancer ; str TILs= stromal tumor-infiltrating lymphocytes ; IT TILs= intratumoral-infiltrating lymphocytes ; NAC=neoadjuvant chemotherapy ; AC=anthracyclines  ; pCR=Pathologic complete response.

**Table S3:** Studies on PD-L1 expression in BC RD

Abbreviations: breast cancer (BC), triple-negative breast cancer (TNBC), hazard ratio (HR), confidence interval (CI), not available (NA), relative risk (RR)

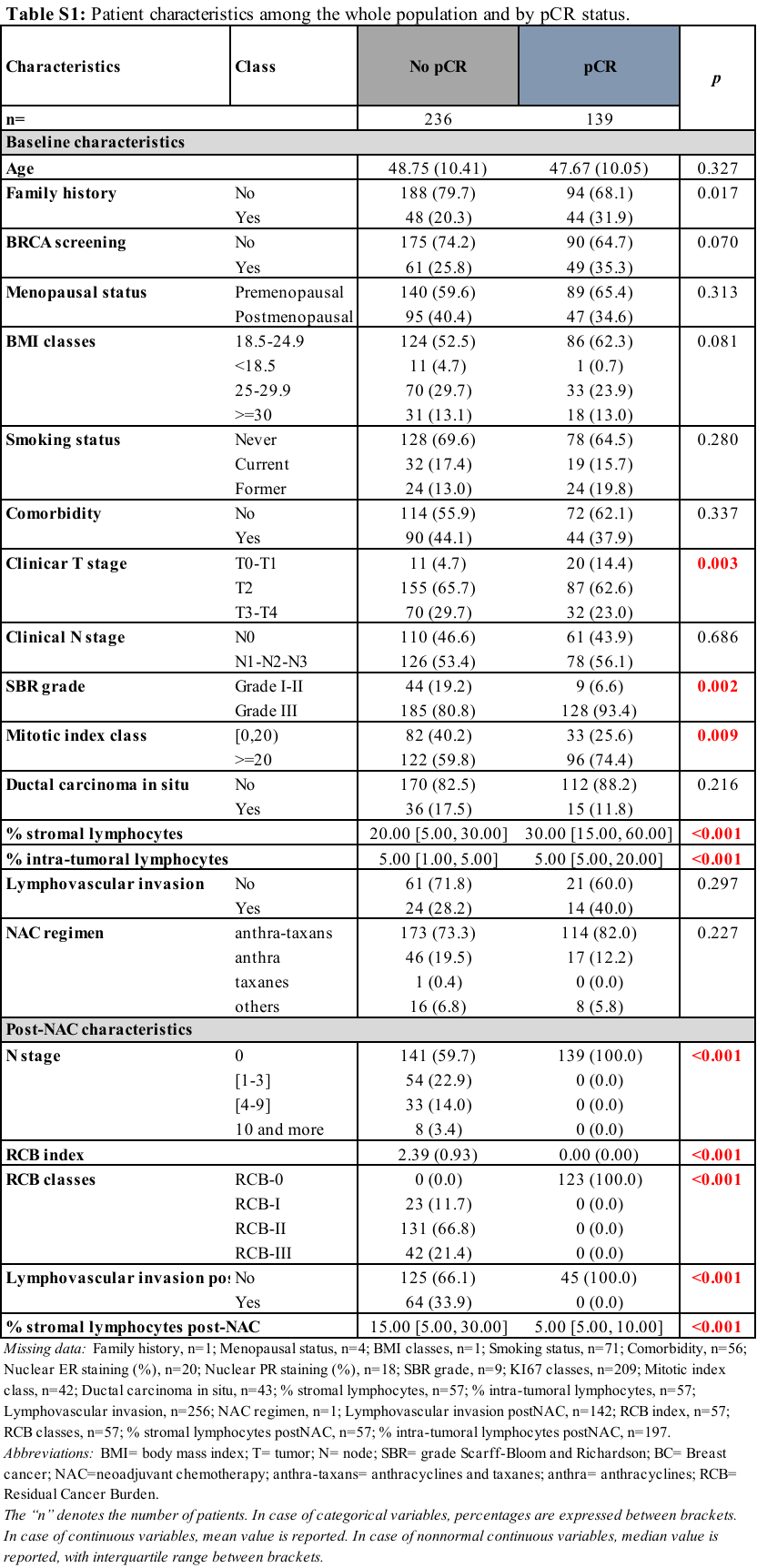

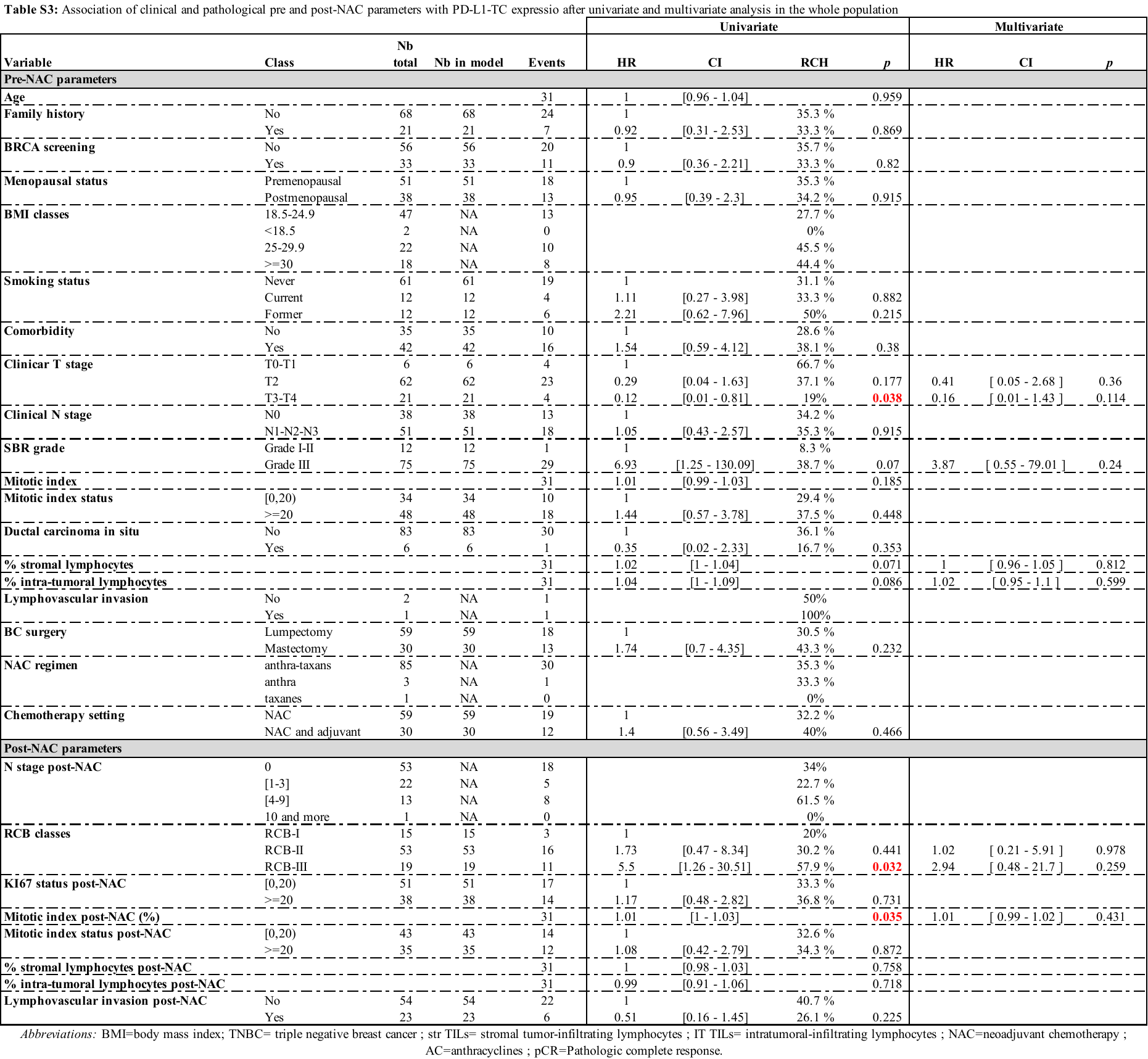

| **First author** | **Journal** | **Year** | **Country** | **n  clinical studies** | **n patients** | **PDL-1 prevalence** | **Association with clinicopathological characteristics** | **PDL1 and pCR** | **PDL1 and DFS** | **PDL1 and OS** |
| --- | --- | --- | --- | --- | --- | --- | --- | --- | --- | --- |
| Wang | The Breast  Journal | 2017 | China | 9 | 8583 | 25.8% (2141/8288) | Ductal BC (p=0.037) High tumor grade (p=0.0001) ER negative (p=0.0001) PR negative (p=0.0001) Her2+ (p=0.001) Basal like (p=0.0001) | - | HR = 1.122 95% CI [0.878; 1.434] p = 0.357 | HR 1.573 95% CI [1.01; 2.45] p = 0.045 |
| Kim | BMC Cancer | 2017 | Korea | 6 | 7877 (827 TNBC) | - | Tumor and immune cells:  Lymph node metastasis (p=0.06) High tumor grade (p<0.001) | - | HR = 1.36 95% CI [1.03; 1.79]  p = 0.03 | HR = 1.908  95% CI [0.91; 4.00] p = 0.09 |
| Zhang | Oncotarget | 2017 | China | 5 | 2546 | 21.7% to 56.6% | Lymph node metastasis (p=0.02) High tumor grade (p<0.001) ER negative (p=0.008) TNBC (p<0.001) | - | - | HR=1.76 95% CI [1.09; 2.82]  p=0.02 |
| Huang | Breast Cancer  Research  and Treatment | 2019 | China | 41 | 14367 | - | Ductal BC (p=0.03) Large tumor size (p=0.05) High tumor grade (p<0.00001) ER negative (p=0.0009) PR negative (p=004) High Ki67 (p=0.01) TNBC (p=0.0001) High TILS (p<0.00001) | Tumor c. : RR = 1.64 95% CI [0.99–2.73] p = 0.05 (n=1085) | Tumor c. :  HR = 1.43 95% CI [1.21; 1.70] p < 0.0001 (n=4349) Immune c. : HR = 0.45  95% CI [0.28; 0.73] p = 0.001 | Tumor c. : HR = 1.58 95% CI [1.14; 2.20] p = 0.006 (n=5890) Immune c. : HR = 0.41  95% CI [0.27; 0.63]  p < 0.0001 |
| Li | Medicine | 2019 | China | 19 | 12505 | 6.4% to 76.4% | Lymph node metastasis (p<0.001) High tumor grade (p<0.001) Negative HR (p<0.001) High Ki67 (p<0.001) High TILS (p<0.001) | - | HR = 1.31 CI95% [1.14; 1.51]  p = 0.0002 | HR = 1.52 CI95% [1.14;2.03] p=0.004 |
| Matikas | Clinical Cancer  Research | 2019 | Sweden | 38 | 9500 | In the TNBC subgroup: tumor cells 41% (n =2740) immune c. 48% (n=1313) tumor and immune c. 46% (n=903) | - | - | Tumor c. overall : HR 1.62  95% CI [1.14–2.33] p=0.008 Immune c. TNBC :  HR =0.61 95% CI [0.51–0.73] p < 0.001 (n=969) | Tumor c. overall :  HR = 1.93 95% CI [1.20; 3.09] p=0.006 Immune c. TNBC : HR = 0.53 95% CI [0.39; 0.73] p < 0.001 |
| Lotfinejad | Diagnostics | 2020 | Iran | 7 | 1152 | - | No associations | - | HR = 0.65 95% CI [0.37; 1.16] p = 0.15 | HR = 0.56 95% CI [0.32 to 1.01] p = 0.056 |

**Table S3.** Recent meta-analyses of the prognostic value of PD-L1 (IHC) in BC *Abbreviations:* breast cancer (BC), estrogen receptor (ER), progesterone receptor (PR), hormone receptor (HR), triple-negative breast cancer (TNBC), tumor-infiltrating lymphocytes (TILS), cells (c.), hazard ratio (HR), confidence Interval (CI), relative risk (RR), pathological complete response (pCR), overall survival (OS, disease-free survival (DFS)

**Supplementary Figures**

**Fig. S1:** Pre-NAC and post-NAC stromal immune infiltration rates in the whole population and by BRCA status. Waterfall plot representing the variation of TIL levels according to PD-L1-TC and PD-L1-IC; each bar represents one sample, and samples are ranked by increasing order of TIL level change. Paired samples for which no change was observed have been removed from the graph. **A,** by PD-L1_TC. B, by PD-L1-IC.

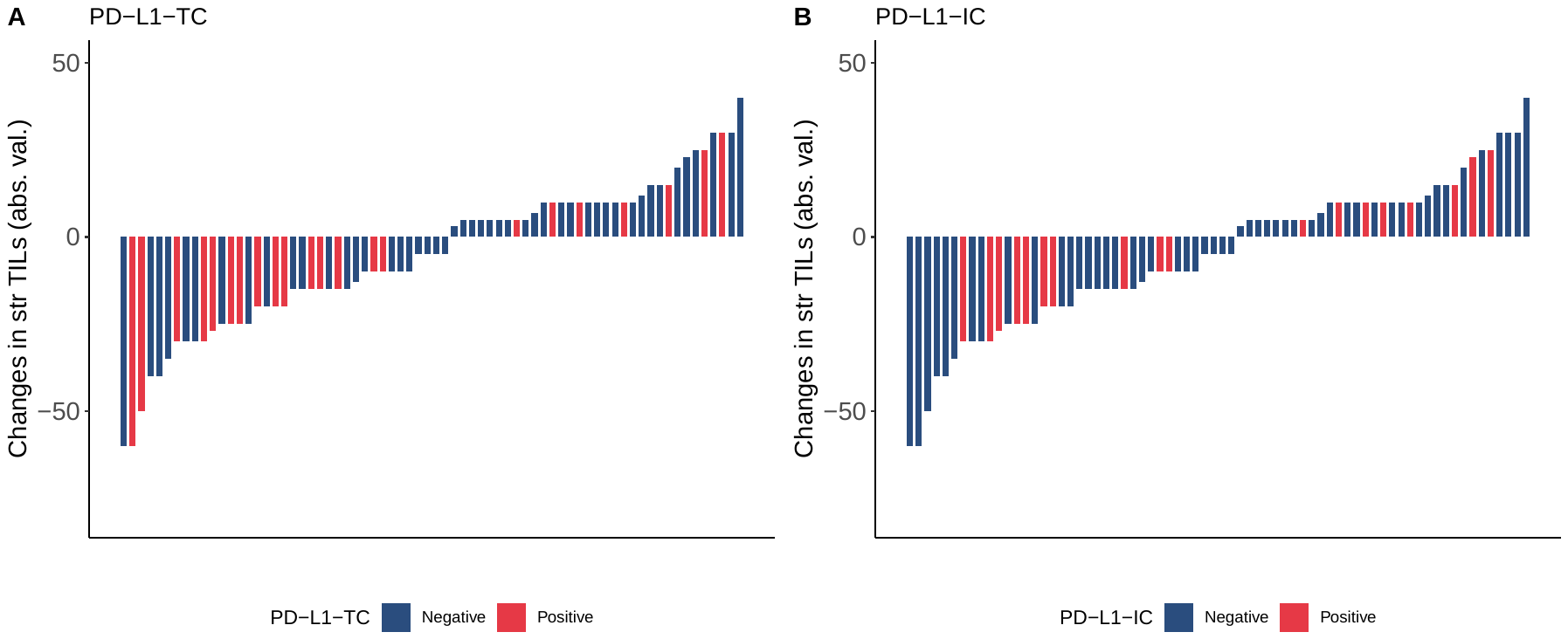
